# Significance of neutrophil-to-lymphocyte ratio, platelet-to-lymphocyte ratio for predicting clinical outcomes in COVID-19

**DOI:** 10.1101/2020.05.04.20090431

**Authors:** Shaoping Huang, Min Liu, Xiaolu Li, Zhiyin Shang, Ting Zhang, Hongzhou Lu

## Abstract

**Background:** The epidemic of 2019 novel coronavirus (COVID-19) struck China in late December, 2019, resulting in about 200000 deaths all over the world. Numerous observational studies have suggested that the neutrophil-to-lymphocyte ratio (NLR) and lymphocyte proportion and the platelet-to-lymphocyte ratio (PLR) are inflammatory markers. Our study aimed to detect the role of NLR, PLR in predicting the prognosis of COVID-19.

**Methods:** Four hundred and fifteen consecutive patients were enrolled in Shanghai Public Health Clinical Center affiliated to Fudan University, between 20 January and 11 April 2020 with confirmed COVID-19. Epidemiology, symptoms, signs, and laboratory examinations during the hospital stay were collected and compared between non-severe and severe patients. Statistical analysis was performed by SPSS 25.0 software.

**Results:** Four hundred and fifteen laboratory-confirmed COVID-19 patients were included in our study, among which 386 (93%) patients were not severe, and 27 (7%) were severe. The proportion of males in severe cases is higher than in non-severe cases (75.86% vs. 50.52%, P=0.008). The age between the two groups is different (p=0.022). Compared with non-severe patients, severe patients exhibited more comorbidities, including hypertension (48.28% vs. 19.43%, p<0.001), diabetes (20.69% vs. 6.99%, p=0.009), chronic obstructive pulmonary disease (51.72% vs. 6.22%, p<0.001), and fatty liver (37.93% vs. 15.8%, p=0.002), respectively. NLR and PLR showed significant difference (p<0.001). Diabetes (OR 0.28; 95% CI 15.824-187.186), fatty liver (OR 21.469; 95% CI 2.306-199.872), coronary heart disease (OR 18.157; 95% CI 2.085-158.083), NLR (OR 1.729; 95% CI 1.050-2.847) were significantly associated with severe cases with COVID-19. The NLR of patients in severe group had a 1.729-fold higher than that of no-severe group (OR 1.729; 95% CI 1.050-2.847, P=0.031).

**Conclusions:** NLR is an independent risk factor of severe COVID-19 patients. PLR, NLR were significantly different between severe and non-severe patients, so assessment of NLR, PLR may help identify high risk cases with COVID-19.

## Introduction

The epidemic of 2019 novel coronavirus (COVID-19) struck China in late December, 2019. A novel coronavirus was then identified as the causative agent. Many countries and territories have been affected within two months[1-3]. As of April 26, 2020, nearly 3,000,000 confirmed cases of COVID-19 occurred, resulting in about 200000 deaths. Many cases were mild to moderate with common symptoms at onset of illness, including fever, cough, and fatigue or myalgia. Organ dysfunction included acute respiratory distress syndrome (ARDS), acute liver injury, acute cardiac injury, acute kidney injury, and death could occur in the severe cases[4]. In our study, we focused on patients with COVID-19 in Shanghai, describing the clinical characteristics and prognostic factors.

Blood cell interactions are essential in the pathophysiology of inflammation, immune responses, hemostasis, and oncogenesis. Numerous observational studies have suggested that the neutrophil-to-lymphocyte ratio (NLR), lymphocyte-to monocyte ratio (LMR), lymphocyte proportion and the platelet-to-lymphocyte ratio (PLR) are inflammatory markers of immune-mediated, metabolic, prothrombotic, and neoplastic diseases, and are widely investigated as useful predictors for prognosis in many diseases[5-7]. These interactions are multifaceted, and it is often difficult to distinguish primary triggering signals and the specific roles of each cell type in the development and progression of disease states. Recent researches of COVID-19 indicated severe patients tended to have higher NLR[8, 9]. Our study aimed to detect the role of NLR, PLR, LMR in predicting the prognosis of COVID-19.

## Material and Methods

### 1. Study Cohort and Design

We recruited 415 consecutive patients in Shanghai Public Health Clinical Center affiliated to Fudan University (the designated hospital in the Shanghai area), between 20 January and 11 April 2020 with confirmed COVID-19. Epidemiology, symptoms, signs, laboratory examinations, chest computerized tomography (CT) scan, and treatment options during the hospital stay were all collected and compared between non-severe and severe patients. This study was approved by the Medical Ethics Committee of Shanghai Public Health Clinical Center affiliated to Fudan University.

### 2. Statistical Analysis

Statistical analysis was performed by SPSS 25.0 software. All data of the demographic and clinical characteristics were expressed as frequencies and proportions for categorical variables, mean SD or median and interquartile (IQR) for continuous variables. We used the one-way ANOVAfor categorical variables and the Mann-Whitney U test for continuous variables. To look at risk factors of severe COVID-19, laboratory values were compared with non-severe and severe patients. Then binary logistic regression analysis was used to analyze the risk factors. A P value of <0.05 was considered to be statistically significant.

## Results

### 1. Demographic and Epidemiological features

415 laboratory-confirmed COVID-19 patients were enrolled in our study, among which 386 (93%) patients were not severe, and 27 (7%) were severe. The demographic, and epidemiologic features on admission were shown in Table 1. Most patients were ethnic Chinese (393/415, 94.7%). The median age of patients was 44 years (IQR 30-61), ranged from 15 years to 84 years. About a half (217, 52.3%) of all patients were male. 220 (53%) patients developed COVID-19 due to contacting with Wuhan, Hubei Province, and 27(6.5%) were imported cases.

**Table 1.** Demographic and Epidemiologic Characteristics of Patients with COVID-19

| Characteristics | All patients,<br>(n=415) | Patients with non-severe<br>COVID-19 (n=386) | Patients with severe<br>COVID-19 (n=29) |
| --- | --- | --- | --- |
| **Demographic** |  |  |  |
| Median age (range), y | 44(30-61) | 41(30-59) | 64.5(62.25-74.75) |
| Men/women, n/n | 217/198 | 195/191 | 22/7 |
| BMI | 23.61(21.24-25.95) | 23.51(21.13-25.95) | 24.22(22.45-27.54) |
| **Ethnicity, n** |  |  |  |
| Ethnic Chinese | 393 | 364 | 29 |
| Non-Chinese | 22 | 22 | 0 |
| **Recent travel to areas with COVID-19**<br>**outbreak, n** |  |  |  |
| Hubei Province | 220 | 204 | 16 |
| Wenzhou, Zhejiang Province | 2 | 2 | 0 |
| Outside China | 27 | 26 | 1 |

### 2. Clinical characteristics

As showed in Table 2, the proportion of males in severe cases is higher than in non-severe cases (75.86% vs. 50.52%, P=0.008). The age between the two groups is different (p=0.022). Compared with non-severe patients, severe patients exhibited more comorbidities, including hypertension (48.28% vs. 19.43%, p<0.001), diabetes (20.69% vs. 6.99%, p=0.009), chronic obstructive pulmonary disease (51.72% vs. 6.22%, p<0.001), and fatty liver (37.93% vs. 15.8%, p=0.002), respectively. Only one patient had HIV infection. 22(5.3%) patients had HBV infection and no significant difference was found between non-severe and severe cases. Among all these cases, 95.34% of the non-severe group were not smokers, which is similar with severe group of 89.66%. Only 3.11% of non-severe cases had drinking history, and the severe cases of 6.9%. For the virus clearance, there were 15 patients (3.89%) showed that the intestinal clearance is later than that of the respiratory.

**Table 2.**
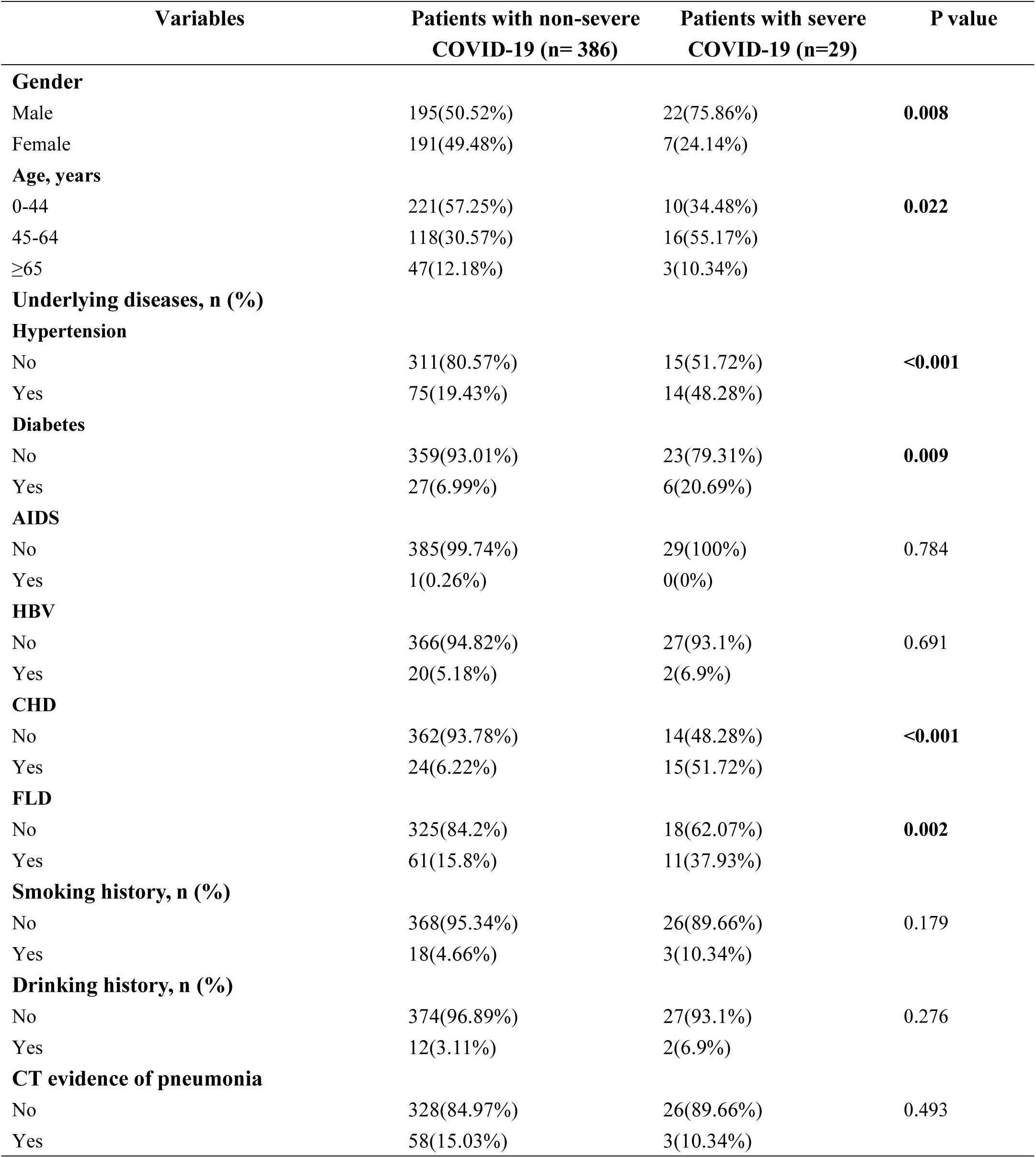
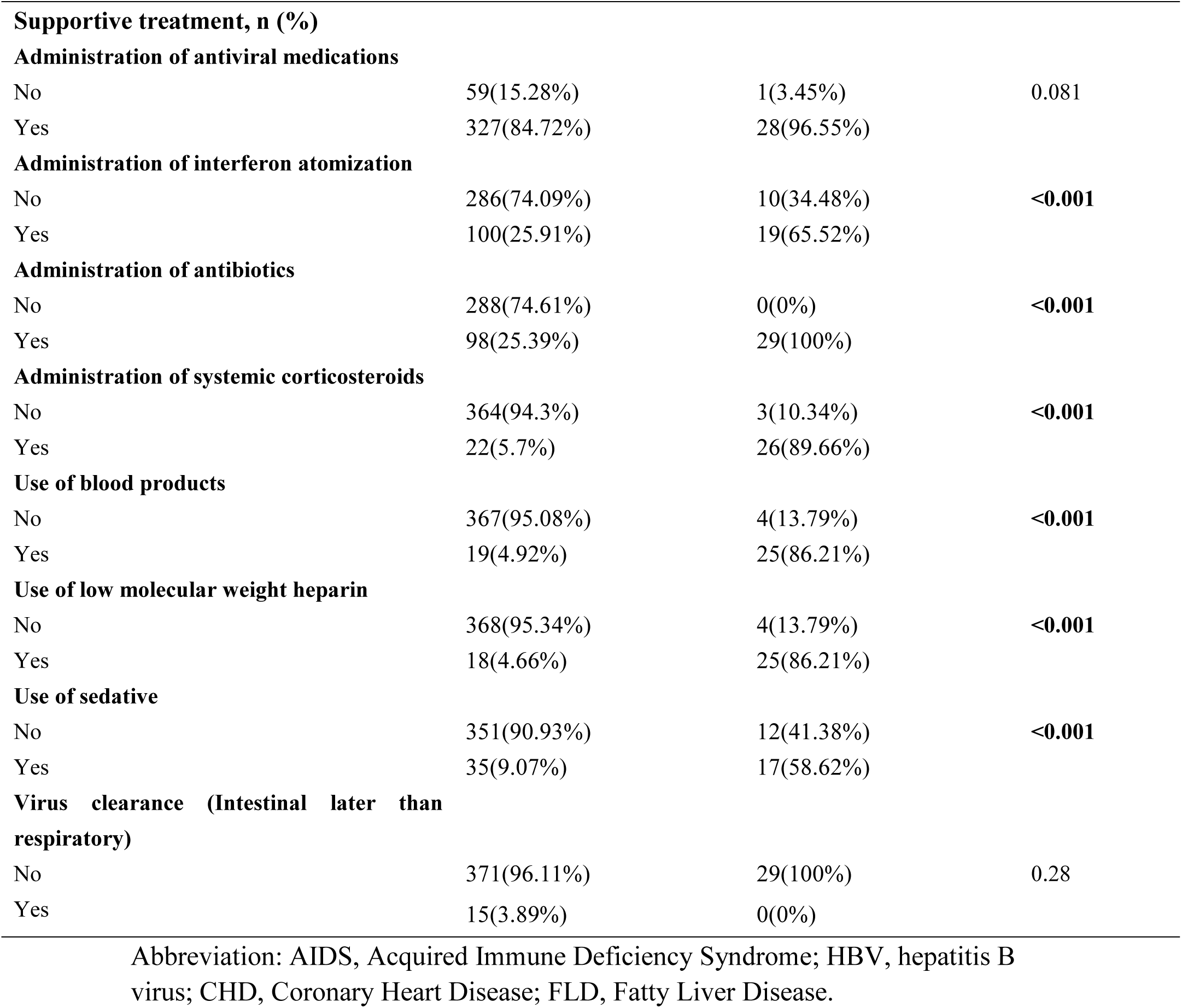
Clinical Features of Patients with COVID-19

| Variables | Patients with non-severe<br>COVID-19 (n= 386) | Patients with severe<br>COVID-19 (n=29) | P value |
| --- | --- | --- | --- |
| **Gender** |  |  |  |
| Male | 195(50.52%) | 22(75.86%) | **0.008** |
| Female | 191(49.48%) | 7(24.14%) |  |
| **Age, years** |  |  |  |
| 0-44 | 221(57.25%) | 10(34.48%) | **0.022** |
| 45-64 | 118(30.57%) | 16(55.17%) |  |
| ≥65 | 47(12.18%) | 3(10.34%) |  |
| **Underlying diseases, n (%)** |  |  |  |
| **Hypertension** |  |  |  |
| No | 311(80.57%) | 15(51.72%) | **<0.001** |
| Yes | 75(19.43%) | 14(48.28%) |  |
| **Diabetes** |  |  |  |
| No | 359(93.01%) | 23(79.31%) | **0.009** |
| Yes | 27(6.99%) | 6(20.69%) |  |
| **AIDS** |  |  |  |
| No | 385(99.74%) | 29(100%) | 0.784 |
| Yes | 1(0.26%) | 0(0%) |  |
| **HBV** |  |  |  |
| No | 366(94.82%) | 27(93.1%) | 0.691 |
| Yes | 20(5.18%) | 2(6.9%) |  |
| **CHD** |  |  |  |
| No | 362(93.78%) | 14(48.28%) | **<0.001** |
| Yes | 24(6.22%) | 15(51.72%) |  |
| **FLD** |  |  |  |
| No | 325(84.2%) | 18(62.07%) | **0.002** |
| Yes | 61(15.8%) | 11(37.93%) |  |
| **Smoking history, n (%)** |  |  |  |
| No | 368(95.34%) | 26(89.66%) | 0.179 |
| Yes | 18(4.66%) | 3(10.34%) |  |
| **Drinking history, n (%)** |  |  |  |
| No | 374(96.89%) | 27(93.1%) | 0.276 |
| Yes | 12(3.11%) | 2(6.9%) |  |
| **CT evidence of pneumonia** |  |  |  |
| No | 328(84.97%) | 26(89.66%) | 0.493 |
| Yes | 58(15.03%) | 3(10.34%) |  |
| **Supportive treatment, n (%)** |  |  |  |
| **Administration of antiviral medications** |  |  |  |
| No | 59(15.28%) | 1(3.45%) | 0.081 |
| Yes | 327(84.72%) | 28(96.55%) |  |
| **Administration of interferon atomization** |  |  |  |
| No | 286(74.09%) | 10(34.48%) | <0.001 |
| Yes | 100(25.91%) | 19(65.52%) |  |
| **Administration of antibiotics** |  |  |  |
| No | 288(74.61%) | 0(0%) | <0.001 |
| Yes | 98(25.39%) | 29(100%) |  |
| **Administration of systemic corticosteroids** |  |  |  |
| No | 364(94.3%) | 3(10.34%) | <0.001 |
| Yes | 22(5.7%) | 26(89.66%) |  |
| **Use of blood products** |  |  |  |
| No | 367(95.08%) | 4(13.79%) | <0.001 |
| Yes | 19(4.92%) | 25(86.21%) |  |
| **Use of low molecular weight heparin** |  |  |  |
| No | 368(95.34%) | 4(13.79%) | <0.001 |
| Yes | 18(4.66%) | 25(86.21%) |  |
| **Use of sedative** |  |  |  |
| No | 351(90.93%) | 12(41.38%) | <0.001 |
| Yes | 35(9.07%) | 17(58.62%) |  |
| **Virus clearance (Intestinal later than respiratory)** |  |  |  |
| No | 371(96.11%) | 29(100%) | 0.28 |
| Yes | 15(3.89%) | 0(0%) |  |

Abbreviation: AIDS, Acquired Immune Deficiency Syndrome; HBV, hepatitis B virus; CHD, Coronary Heart Disease; FLD, Fatty Liver Disease.

Patients with COVID-19 were treated with different drugs. From our data, patients with severe COVID-19 treated with antiviral medications, interferon atomization, antibiotics, blood product, systemic corticosteroids, and low molecular weight heparin, which were significantly higher rate than that in the patients with no-severe COVID-19 (P<0.05).

### 3. Laboratory Findings

The laboratory findings were summarized in Table 3 and Table 4. Many patients had hematologic abnormalities on admission: anemia (hemoglobin level< 120 g/L) in 13.5% of patients, leukopenia (leukocyte count <4 ×10^9^cells/L) in 23.9%, lymphopenia (lymphocyte count<1 ×10^9^ cells/L) in 38.8%, and high monocyte percentage (>10%) in 11.5%. The prothrombin time was prolonged (>14 seconds) in the 5.8% of patients. Alanine aminotransferase (>40U/L) were exhibited in 17.5% of the patients, and 54.3% of the patients had elevated C-reactive protein levels (>6 mg/L).

**Table 3.** Laboratory Findings of Patients with COVID-19

| Laboratory Finding | Patients with non-severe<br>COVID-19(n=321) | Patients with severe<br>COVID-19(n=27) | P value |
| --- | --- | --- | --- |
| Leukocyte count, $\times 10^9$ cells/L | 4.68(3.73-5.66) | 6.24(4.47-7.74) | 0.082 |
| Neutrophil, % | 63.29±11.44 | 72.48±13.70 | 0.001 |
| Neutrophil count, $\times 10^9$ cells/L | 2.85(2.28-3.79) | 4.06(3.26-6.42) | **0.004** |
| Lymphocyte, % | 24.50(20.6-31.9) | 15.5(5.9-21.6) | **<0.001** |
| Lymphocyte count, $\times 10^9$ cells/L | 1.12(0.82-1.49) | 0.88(0.45-1.22) | **0.001** |
| Monocyte, % | 9.5(7.3-11.3) | 7.2(4.0-13.6) | **0.015** |
| Monocyte count, $\times 10^9$ cells/L | 0.41(0.35-0.58) | 0.50(0.24-1.07) | 0.157 |
| Hemoglobin level, g/L | 138.29±16.15 | 129.86±29.99 | 0.017 |
| Hematokrit, % | 40.21±4.35 | 38.92±4.20 | 0.143 |
| Platelet count, $\times 10^9$ cells/L | 177(143-220) | 155(117-189.5) | 0.455 |
| C-reactive protein level, mg/L | 6.06(0.499-18.45) | 70.74(31.02-103.77) | **<0.001** |
| ESR, mm/60min | 48(25-89) | 82(42.5-110.5) | **0.027** |
| Prothrombin time, s | 13.5(13.0-13.8) | 13.9(12.85-15.55) | 0.266 |
| FDP | 0.88(0.39-1.47) | 2.42(1.29-20.25) | **0.006** |
| D-dimer | 0.43(0.24-0.57) | 0.89(0.57-5.26) | **<0.001** |
| BNP | 29.41(21.05-46.28) | 133.30(56.38-504.4) | **0.005** |
| Procalcitonin, ng/mL | 0.03(0.02-0.05) | 0.14(0.07-0.35) | **<0.001** |
| Alanine aminotransferase level, U/L | 20(14-34) | 25(14-37) | 0.289 |
| Albumin level, g/L | 41.71±4.59 | 36.61±5.94 | **<0.001** |
| Globulin level, g/L | 28.22(25.9-30.91) | 27.95(25.22-31.35) | 0.06 |
| Creatinine | 64.48(51.73-74.36) | 66.87(56.30-80.93) | 0.149 |
| eGFR | 112.16(95.74-131.56) | 99.70(81.31-117.08) | 0.166 |
| CK-MB | 11.31(9.68-15.13) | 14.42(10.86-20.58) | **0.003** |
| Total bilirubin level, mmol/L [mg/dL] | 8.2(6.3-13.8) | 10.3(6.95-15.0) | 0.049 |
| Lactate, mmol/L | 2.67(2.31-3.07) | 2.50(2.31-3.64) | 0.534 |
| Lactate dehydrogenase level, U/L | 214(190-268) | 399(278-537) | **<0.001** |
| NLR | 2.67(1.76-3.42) | 4.16(3.14-14.65) | **<0.001** |
| LMR | 2.73(1.84-3.66) | 1.92(0.51-3.13) | 0.052 |
| PLR | 145.64(119.87-196.68) | 180.68(110.56-457.96) | **0.004** |

**Table 4.** Laboratory Findings of Patients with COVID-19

| Laboratory Finding | Patients with non-severe<br>COVID-19(n=321) | Patients with severe<br>COVID-19(n=27) | P value |
| --- | --- | --- | --- |
| Lactate dehydrogenase level, U/L |  |  |  |
| ≤290.5 | 315(81.82%) | 10(34.48%) | <0.001 |
| >290.5 | 70(18.18%) | 19(65.52%) |  |
| NLR |  |  |  |
| ≤3.5 | 289(74.87%) | 8(27.59%) | <0.001 |
| >3.5 | 97(25.13%) | 21(72.41%) |  |
| LMR |  |  |  |
| ≤3.0 | 209(54.15%) | 19(65.52%) | 0.236 |
| >3.0 | 177(45.85%) | 10(34.48%) |  |
| PLR |  |  |  |
| ≤222.5 | 318(82.38%) | 15(51.72%) | <0.001 |
| >222.5 | 68(17.62%) | 14(48.28%) |  |

Our study discovered that the white blood cell (WBC) counts on admission of the severe and non-severe cases had no significant difference (P=0.082). Neutrophil counts were significantly higher in severe cases than non-severe cases (P<0.05). Whereas lymphocyte counts were significantly lower in severe cases (0.88× 10^9^/L) than non-severe cases (1.12 × 10^9^/L). The monocyte percentage was also different between the 2 groups (P=0.015). Severe patients had lower hemoglobin level than that of non-severe patients (P=0.017). NLR, LMR and PLR were compared with severe and non-severe cases, among which NLR and PLR were significant different (P<0.001).

Compared with non-severe cases, inflammation-related marker levels (CRP, ESR) were significantly higher in severe cases (P<0.05). The levels of procalcitonin, NT-proB-type natriuretic peptide (NT-proBNP), d-dimer, FDP, albumin, lactate dehydrogenase (LDH), total bilirubin, conjugated bilirubin, CK-MB were elevated in all patients, and significant difference was found between non-severe and severe groups (P<0.05).

### 4. Analysis with LDH, NLR, LMR and PLR

To further define the value of the inflammatory-related parameters in predicting clinical outcomes in COVID-19 patients, the optimal cut-off values for preoperative LDH, NLR, LMR, and PLR that best predicted were calculated as followed: 290.5 (area under the curve (AUC): 0.728; sensitivity: 65.5%; specificity: 88.8%), 3.5 (AUC: 0.764; sensitivity: 72.4%; specificity: 75.1%), 3.0 (AUC: 0.608; sensitivity: 70.0%; specificity: 63.9%), and 222.5 (AUC: 0.659; sensitivity: 48.3%; specificity: 82.6%), respectively (Fig. 1A-D). Then, the patients were dichotomized into high or low LDH/NLR/PLR/LMR groups with these cut-off values. We found that there were significant differences in LDH, NLR and PLR between severe and non-severe groups. (P<0.001).

**Fig. 1.**
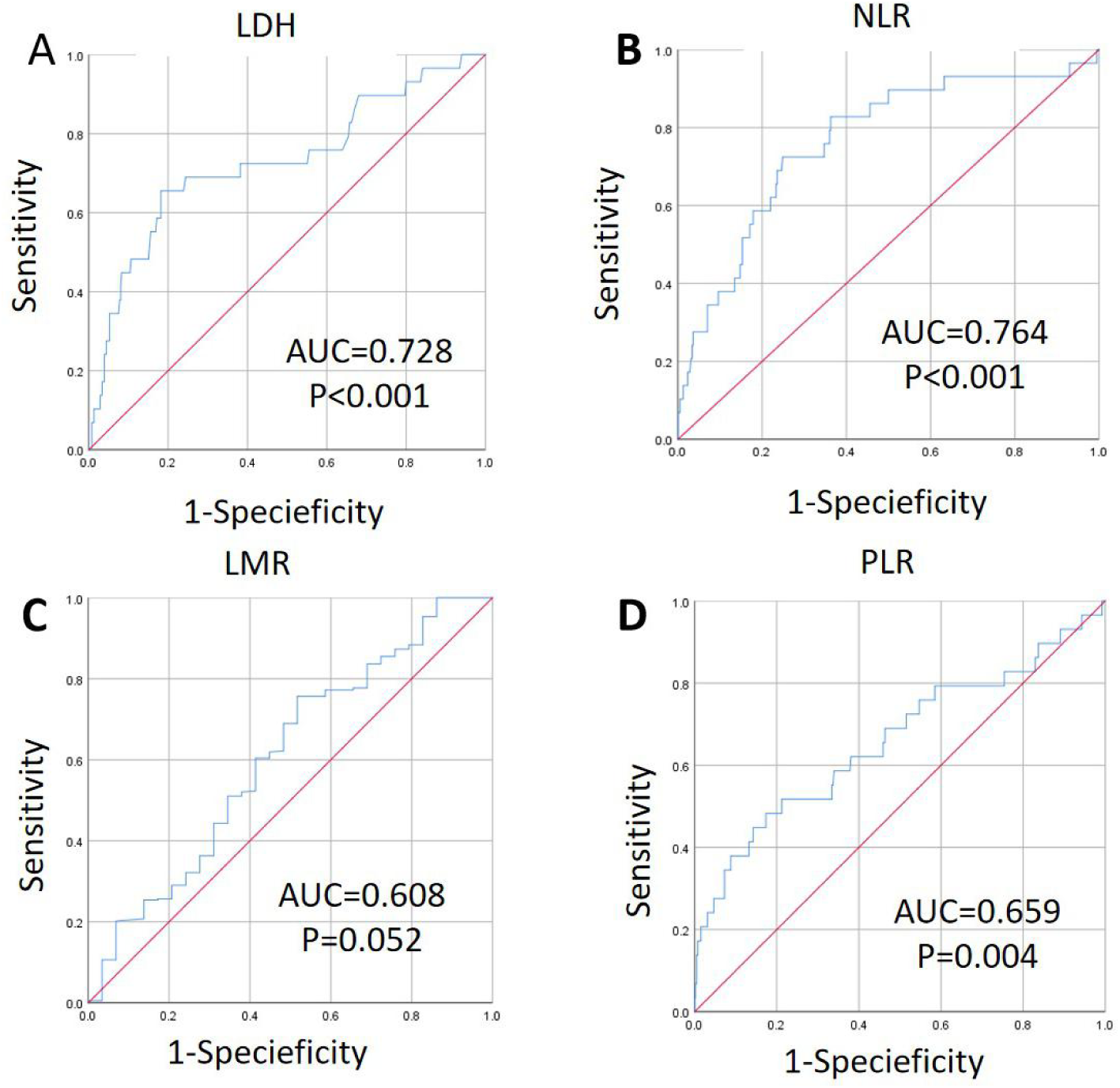
The cut-off values for the inflammation-related parameters. A-D. ROC curves were adopted to calculate the cut-off values for LDH(A), NLR (B), LMR (C), and PLR(D). LDH, lactate dehydrogenase, NLR, neutrophil-to-lymphocyte ratio, LMR, lymphocyte-to-monocyte ratio, PLR, platelet-to-lymphocyte ratio, AUC, area of under curve.

### 5. Risk factors for severe cases

In the logistic regression model, variables such as diabetes (OR 0.28; 95% CI 15.824-187.186), fatty liver (OR 21.469; 95% CI 2.306-199.872), coronary heart disease (OR 18.157; 95% CI 2.085-158.083), NLR (OR 1.729; 95% CI 1.050-2.847) were significantly associated with severe cases with COVID-19, while ESR is a protective factor (OR 0.949; 95% CI 0.940-0.999) for no-severe patients (Table 5). The forecast accuracy is 96.6%.

**Table 5.** Risk factors for severe cases

| Variables | P | OR | 95% CI for OR |  |
| --- | --- | --- | --- | --- |
|  |  |  | Lower | Upper |
| Diabetes | 0.028 | 15.824 | 1.338 | 187.186 |
| FLD | 0.007 | 21.469 | 2.306 | 199.872 |
| CHD | 0.009 | 18.157 | 2.085 | 158.083 |
| NLR | 0.031 | 1.729 | 1.050 | 2.847 |
| ESR | 0.043 | 0.969 | 0.940 | 0.999 |

In order to better diagnose severe COVID-19, we use the ROC curves to calculate the optimal cut-off values for the combined predictive indicator and other risk factors (Figure 2). The combined predictive indicator which is formulated by logistic regression models, is superior to any single indicator in predicting severe patients.

**Fig. 2.**
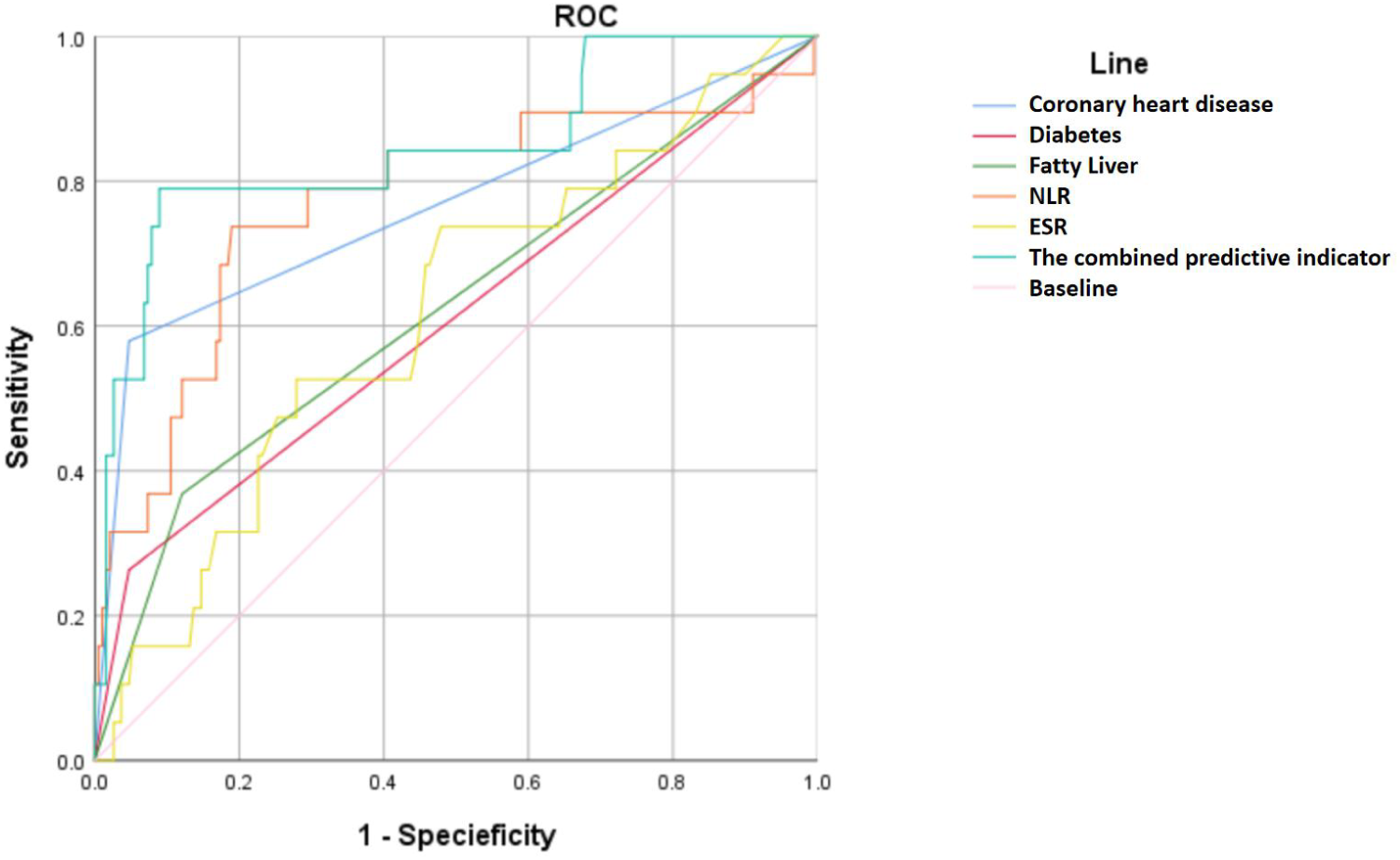
The combined predictive indicator and other risk factors for ROC curves. NLR, neutrophil-to-lymphocyte ratio, ESR, erythrocyte sedimentation rate.

## Discussion

Nowadays, COVID-19 outbreak has caused widespread concern and threatened the global public health security. This retrospective cohort study included 415 COVID-19 patients, among which 29 (7%) were severe. This is different from reports from Wuhan, China [2, 10]. The clinical, laboratory, and radiographic features in this cohort of patients with COVID-19 in Shanghai were nonspecific and similar to those in other series [1-4, 11, 12]. Recently, the epidemics has transmitted from the first stage, in which imported cases composed of the main laboratory-confirmed cases in Shanghai. Compared with patients who had non-severe COVID-19, patients with severe COVID-19 had lower lymphocyte percentage, lymphocyte counts, monocyte percentage, and higher neutrophil percentage, neutrophil counts at presentation. These patients may have had a higher viral load at presentation, which may have led to the apparently worse of laboratory values. We also compared NLR and PLR, the two groups have significantly differences. This probably due to their viral load and immune state. Lactate dehydrogenase, C-reactive protein (CRP), and D-dimer, prothrombin time, FDP, BNP were higher in severe groups than in non-severe groups. From Guang Chen’s research, they found that the SARS-CoV-2 infection may affect primarily T lymphocytes particularly CD4+T and CD8+ T cells, resulting in decrease in all numbers as well as IFN-γ production[11]. Our data analyzed these immune markers in order to find the cause of severity of COVID-19.

In this cohort, we observed that 66.0% of the patients had at least one underlying disorder (i.e., hypertension, diabetes, coronary heart disease), and a higher percentage of hypertension and fatty liver in the severe cases than the non-severe individuals, in consistent with other studies [12, 13]. The risk factors for severity included age, high LDH level, and high d-dimer level in previous reports [8]. However, different from the findings of previous studies, we found coronary heart disease and fatty liver were the comorbidity associated with the severity of COVID-19.

The NLR was higher in severe cases than in non-severe cases which is consistent with recent studies [8, 9]. Besides, we found PLR was also higher in severe patients compared with non-severe patients. Numerous observational studies have suggested that the NLR, LMR, lymphocyte proportion and the PLR are inflammatory markers of immune-mediated, metabolic, prothrombotic, and neoplastic diseases, and are widely investigated as useful predictors for prognosis in many diseases [5-7]. The results of this study have several clinical implications and strengths. Since NLR and PLR could be quickly calculated based on a blood routine test on admission, so we should pay attention to these laboratory findings to identify high risk COVID-19 patients.

Recent researches focus on asymptomatic infection [14, 15], possible fecal-oral transmission in SARS-Cov-2 infection [16], and positive result for SARS-Cov-2 test in recovered patients [17]. At the same time, some reports discussed some COVID-19 patients with underlying disorders, such as diabetes [18], cancer [19] and so on. As we all know, people around the world should pay attention to this disease.

There are some limitations in our study. First, the number of observed events is to some extent small which may limit the statistical power of this research. However, the sample size is sufficient to draw a conclusion. Second, in our group, there were fewer severe patients which may not balance for analysis. Third, the causal relationship between abnormal laboratory findings and severity could not be estimated since laboratory findings were measured on admission and may not indicate the severity of COVID-19.

## Data Availability

This article is available.

